# Hematologist-level classification of mature B-cell neoplasm using deep learning on multiparameter flow cytometry data

**DOI:** 10.1101/2020.03.31.20041442

**Authors:** Max Zhao, Nanditha Mallesh, Richard Schabath, Alexander Höllein, Claudia Haferlach, Torsten Haferlach, Franz Elsner□, Hannes Lüling□, Peter Krawitz, Wolfgang Kern

## Abstract

The wealth of information captured by multiparameter flow cytometry (MFC) can be analyzed by recent methods of computer vision when represented as a single image file. We therefore transformed MFC raw data into a multicolor 2D image by a self-organizing map (SOM) and classified this representation using a convolutional neural network (CNN). By this means, we built an artificial intelligence that is not only able to distinguish diseased from healthy samples, but that can also differentiate seven subtypes of mature B-cell neoplasm (B-NHL). We trained our model with 18,274 cases including chronic lymphocytic leukemia (CLL) and its precursor monoclonal B-cell lymphocytosis (MBL), marginal zone lymphoma (MZL), mantle cell lymphoma (MCL), prolymphocytic leukemia (PL), follicular lymphoma (FL), hairy cell leukemia (HCL), lymphoplasmacytic lymphoma (LPL) and achieved a weighted F1 score of 0.94 on a separate test set of 2,348 cases. Furthermore, we estimated the trustworthiness of a classification and could classify 70% of all cases with a confidence of 0.95 and higher. Our performance analyses indicate that particularly for rare subtypes further improvement can be expected when even more samples are available for training.

## Introduction

The number of markers in flow cytometry has increased considerably over the recent years. However, the strategy to analyze multiparameter flow cytometry (MFC) data by human experts remained essentially the same: cell populations of interest are identified by a sequential gating procedure in 2D scatter plots. Although several computational methods have been developed that were able to reach manual gating accuracies (1,2), these approaches cannot be used for differentiating multiple subtypes of hematological disorders (2). Recently, deep convolutional neural networks (CNN) have been used successfully for different classification tasks on medical imaging data (3,4). We therefore hypothesized that visualization techniques transforming MFC raw data into a single image file, might be a promising representation for a multi-class problem that can be investigated by a CNN. We therefore extended existing approaches that are based on self-organizing maps (SOM), such as flowSOM (5), to generate an input for a CNN.

A SOM is a network of interconnected nodes or neurons, often ordered in a 2-dimensional topology, that can be used for unsupervised clustering of high-dimensional data (6). In our case, the input to the SOM is the light emission profiles from fluorescent dyes coupled to specific antibodies detecting specific epitopes on the cell surfaces. This input is mapped to the most similar node in the SOM. This node as well as neighboring nodes are updated so that structural properties of the original data are preserved. This means, cells with a similar expression profile will be assigned to the same node or a node close by in the SOM.

The SOM-transformed data can then be analyzed by a CNN, which is a network architecture that is frequently used for pattern recognition in medical image data, such as portrait photos of dysmorphic patients or radiographs (7). However, CNNs require a large number of samples for training to achieve good performance (8,9). We therefore chose the following seven mature B-cell neoplasm (B-NHL) subtypes for classification, as a data set of more than 20,000 samples was available, that was gathered using the same flow cytometry protocol: chronic lymphocytic leukemia (CLL) and its precursor monoclonal B-cell lymphocytosis (MBL), marginal zone lymphoma (MZL), mantle cell lymphoma (MCL), prolymphocytic leukemia (PL), follicular lymphoma (FL), hairy cell leukemia (HCL), and lymphoplasmacytic lymphoma (LPL).

## Methods

### Flow cytometry data

MFC data (fcs files) was obtained from 20,622 routine diagnostic samples from patients with suspected B-NHL that had been analyzed between 2016-01-01 and 2018-12-31 at MLL Munich Leukemia Laboratory including normal controls and eight different B-cell lymphoma subtypes distributed as follows: CLL (n=4,438), PL (n=588), FL, (n=246), HCL (n=225), LPL (n=726), MCL (n=313), MZL (n=1,106), MBL (n=1,614), and healthy controls (n=11,366). 2,348 samples that were labeled after the date of 2018-07-01 were not used for training or validating the model, but exclusively for testing. Unlike randomly splitting data into training and validation sets and testing sets, preserving chronological order represents a more realistic situation.

Validation data is taken from a random split of the training data collected before 2018-07-01. Detailed number of samples in training and test set can be found in Table 1. For the assessment of B-NHL, a panel consisting of three 9-color combinations of monoclonal antibodies was used in all samples to analyze the surface expression of 21 antigens with detailed antibody-color combinations given in Supplement Table 1. In the following we refer to this panel as B-NHL panel. A target of 50,000 events were acquired per analysis on a Navios cytometer (Beckman Coulter, Miami, FL). We also measured 279 additional samples with acute myeloid leukemia (AML, n=124), multiple myeloma (MM, n=101) and hairy cell leukemia variant (HCLv, n=54) with the B-NHL panel, in order to assess the behavior of preprocessing and classification on previously untrained B-NHL subtypes and non B-NHL hematological disorders. Only samples obtained from peripheral blood or bone marrow aspirate were included in the analysis. Flow cytometry data is stored in FCS 2.0 format (10). PnG-scaling (linear gain with a factor) was used for SS and FS and PnE-scaling (exponential scaling) for all color channels. All samples were ensured of having identical PnG and PnE parameters. No re-linearization or additional transformation were applied to the data. Visual inspection of the channel plots was performed to detect issues in machine calibration and compensation in the dataset. Single channel intensity plots are shown in Supplement Figure 1. For further processing in each sample, all channels were normalized by rescaling their minimum and maximum to the range of 0 to 1 without changing the initial distribution.

**Table 1.** Included B-cell lymphoma cohorts

| | CLL | MBL | MCL | PL | LPL | MZL | FL | HCL | normal | $\Sigma$ |
| --- | --- | --- | --- | --- | --- | --- | --- | --- | --- | --- |
| Training data | 4035 | 1554 | 275 | 552 | 642 | 990 | 222 | 201 | 9803 | 18274 |
| Test data | 403 | 60 | 38 | 36 | 84 | 116 | 24 | 24 | 1563 | 2348 |

*Note:* Data collected of patients with suspected lymphoma between January and December 2018 amounted to a total of 20622 individual patients. Diagnoses both with conclusive and inconclusive results in flow cytometry were included, as long as they were clearly diagnosed for a single B-cell lymphoma. Patients with multiple hematological neoplasia were not included in the assessment. The training data is used for all model development and model comparisons, as well as validation data. The final test dataset includes all patients after a time deadline of 01.04.2018.

**Figure 1.**
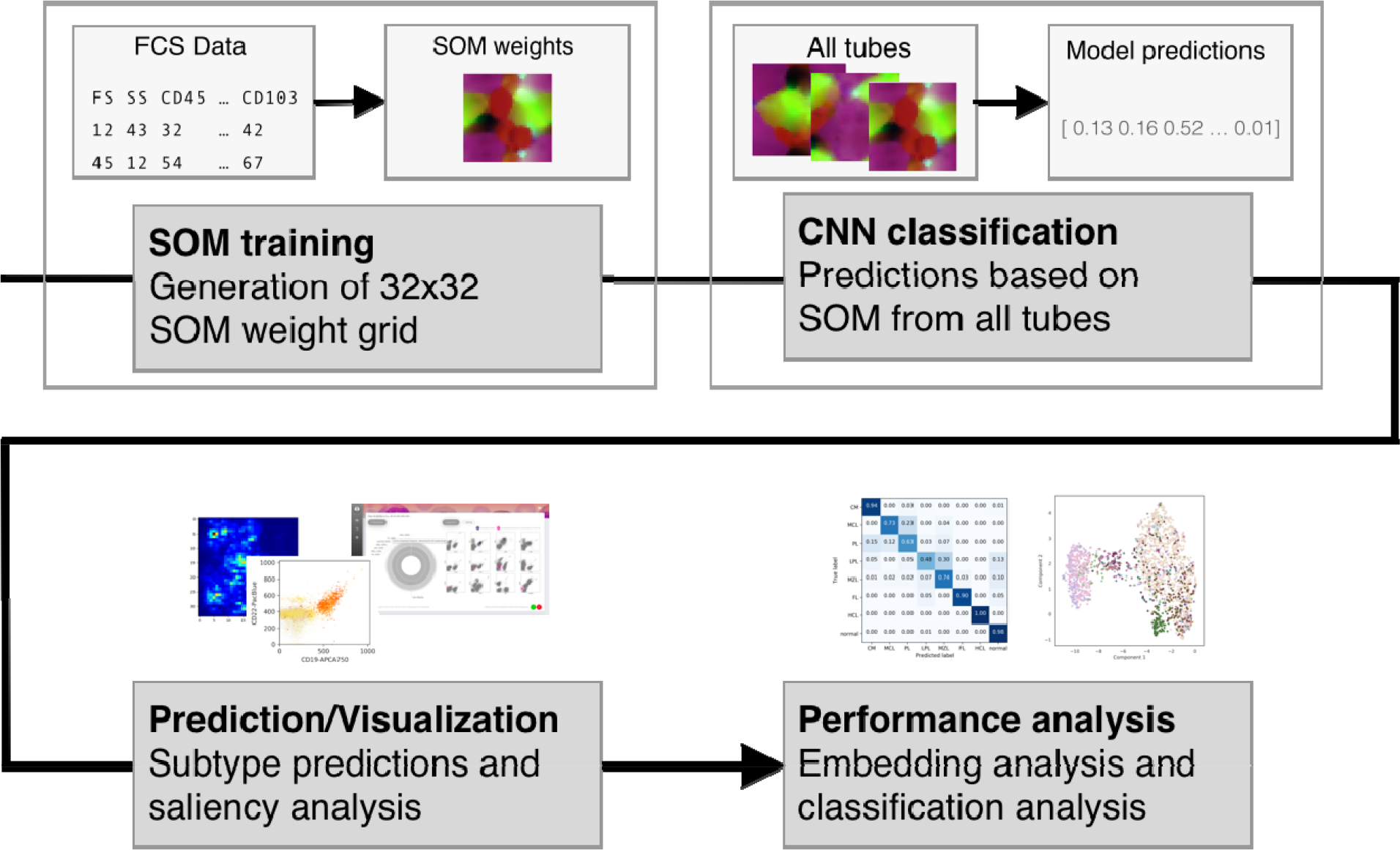
Overview of the classification pipeline. Individual 2D SOMs are generated for each tube of a single case. The weights of the SOM nodes are used as input for a CNN that predicts lymphoma subtypes. The trustworthiness of a suggested diagnosis is computed and the cells contributing most to this decision are visualized in density plots and saliency maps. The overall performance of the classification process is benchmarked with a confusion matrix and the similarity of cases is visualized by a t-SNE plot.

### SOM transformation

Prior to classification, SOMs were used as a method that reduces the dimensionality of the data, but that preserves its spatial structure. After training on the MFC data the weights of the SOM were further processed. For each patient, data from individual tubes were independently transformed into separate SOMs. These SOMs were simultaneously used as inputs to a classifier for generating predictions. The SOM model was adapted from an implementation using tensorflow for GPU-based training (11). All MFC events were mapped in batch to calculate updates increasing the throughput of the training algorithm by leveraging efficient vectorizations□(12). The Euclidean norm was used to calculate the distance between input vectors and single SOM nodes. A radius parameter was used to set the width of Euclidean neighborhood function for calculating weight updates. In an initial assessment (Supplement Figure 5) of training parameters showed that a higher learning radius correlated with lower topographic error (TE) higher mean quantization error (MQE). TE describes the proportion of input entries where the first and second best matching nodes are non-adjacent (13). MQE uses the average Euclidean distance of each input to their best matching node. Larger number of epochs only played a minor role in increasing MQE after a certain threshold. The quality of the created SOMs is similar to flowSOM as shown in Supplementary Figure 2.

**Figure 2.**
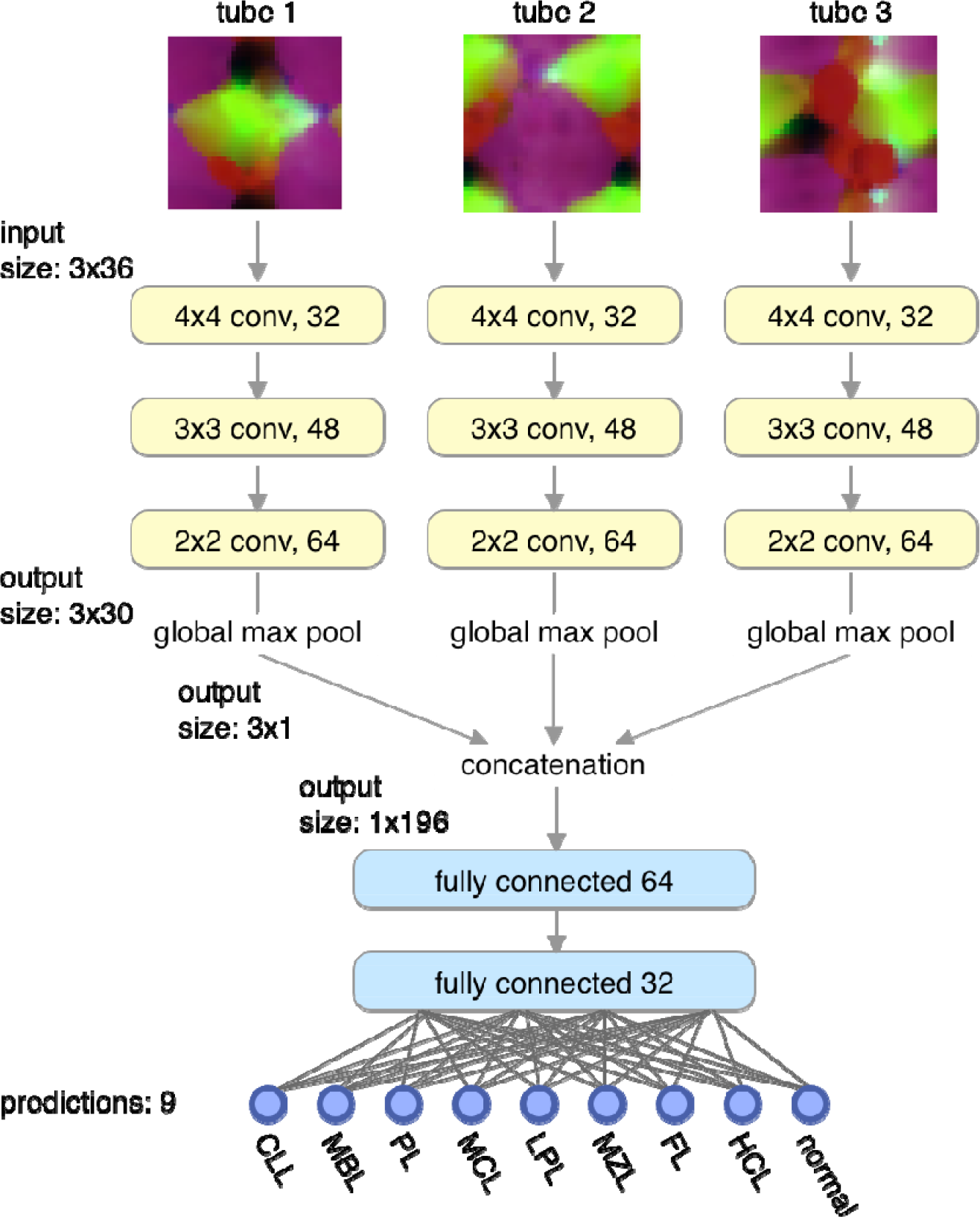
Architecture of the CNN. First, the original 32×32 SOMs are toroidally wrapped by 2 pixels on each edge to produce a 36×36 input matrix, which is fed into convolutional layers with 32 4×4 filters. The input from each SOM is processed individually in a sequence of convolutional layers (conv), followed by a global max pooling and concatenation layer. This vector is further processed in two fully connected hidden layers and results in a softmax prediction layer.

Individual SOM transformation uses preinitialized weights from a reference SOM trained on a small subset of samples. The reference SOM uses a random sample from each diagnostic subtype in the training dataset with at least 20% infiltration reported in the manual analysis. These cases are used to train the reference SOM from random initialization for each tube. Training uses 10 epochs and an initial radius of 24, linearly decreasing to 2 at the end of training. A toroidal neighborhood function was used to avoid edge artifacts caused by a planar map (14). Transformation of the MFC data to individual SOM uses 4 training epochs and a starting radius of 4, linearly decreasing to 1.

### Prediction model

The model architecture and training graphs are shown in Figure 1. The basic model architecture generates predictions using SOM node weights for a number of classes. The weights are initially processed in three convolutional layers with decreasing filter sizes, which is followed by a global max pooling layer, summarizing filters across the spatial dimension of the SOM map. Afterwards two dense layers are used to combine information for class prediction. In order to merge information from multiple tubes, the convolutional layers and a final max pooling layer are replicated for each tube and the result from each max pooling layer is concatenated across all tubes and used in the dense layers. Thus, the dense layers combine information from all provided tubes. Models have been trained for 15 epochs using the Adam optimizer□(15) with a learning rate of 0.001. Global weight decay of 5e-5 has been applied to all layers. The model has been implemented in Keras□(16). Classification accuracies are evaluated on a 10% validation split and was used to optimize the network architecture. The hold-out test set is used to assess the model performance. The CNN model was compared to classical machine learning methods and a simpler dense neural network implementation with a similar number of trainable parameters. The CNN achieved superior overall accuracy and average F1scores in comparison to alternative classifiers. Initial results (Supplement Table 2) for model selection and optimization on the 10% validation data set indicated higher classification performance of the CNN model (F1 0.70) in comparison to dense neural nets (F1 0.61) and classical machine learning classifiers, such as random forest (F1 0.45). Results were generated for 10×10 SOMs. Additionally, 32×32 and 10×10 performance were compared in the CNN model, showing higher classification performance in the former (F1 0.76). 32×32 SOMs were chosen as the base SOM size for all following results, because of better classification performance and only marginal performance penalties in SOM training. Larger SOM sizes such as 48×48 were not attempted because of performance and dimensionality reduction considerations. Results are shown in Supplement Table 2.

### Benchmarking of subtype inference

The class with the highest score was used as the predicted label in analysis. Thresholds were defined, below which results were relegated to an additional *uncertain* class. Precision and recall per group were defined on the true label of each case. Predictions performance was evaluated with average-per-group F1 scores calculated as avg 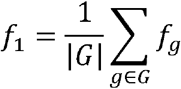, with 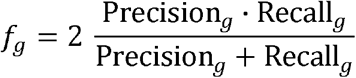 and *G* as the set of all groups.

This can also be viewed as the calculation of F1 scores splitting the predictions into groups with the same true label. Weighted F1 was calculated as the group-size weighted average of per-group F1 scores, which is calculated as weighted 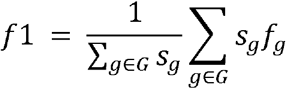 with *s*_*g*_ as the number of cases in Group *g*.

t-SNE was used to visualize embedding pattern of classes on flattened input data. Parameters of a perplexity of 50, a learning rate of 200 and 1000 iterations were chosen for t-SNE fitting. Results for intermediate layer are presented in Figure 4b.

### Channel importance analysis via data occlusion

Importance of individual MFC channels for prediction accuracy in the trained model was measured by zeroing all values in the respective channel in the input SOM, which we will refer to as occlusion analysis. The approach has been previously described in model analysis of image classifiers (17). More important information in the original input data will decrease prediction accuracy more strongly and thus increase the measured loss. Important channels for each group are calculated using average cross-entropies in each group for all channels.

### Visualization of informative cell populations

The entire area of a scatter plot can be subdivided into small sections and cells can be assigned accordingly. By this means we computed, how densely each part of the scatter plot is usually populated by cells of a certain diagnostic subtype. The density maps of the subtypes can be compared to healthy samples and areas that are under- or overpopulated can be indicated by color (Fig. 3). When cells of a single sample are inspected in the scatter plot, this color scheme helps to interpret whether a cell cluster is representative for a certain diagnosis or not.

**Figure 3.**
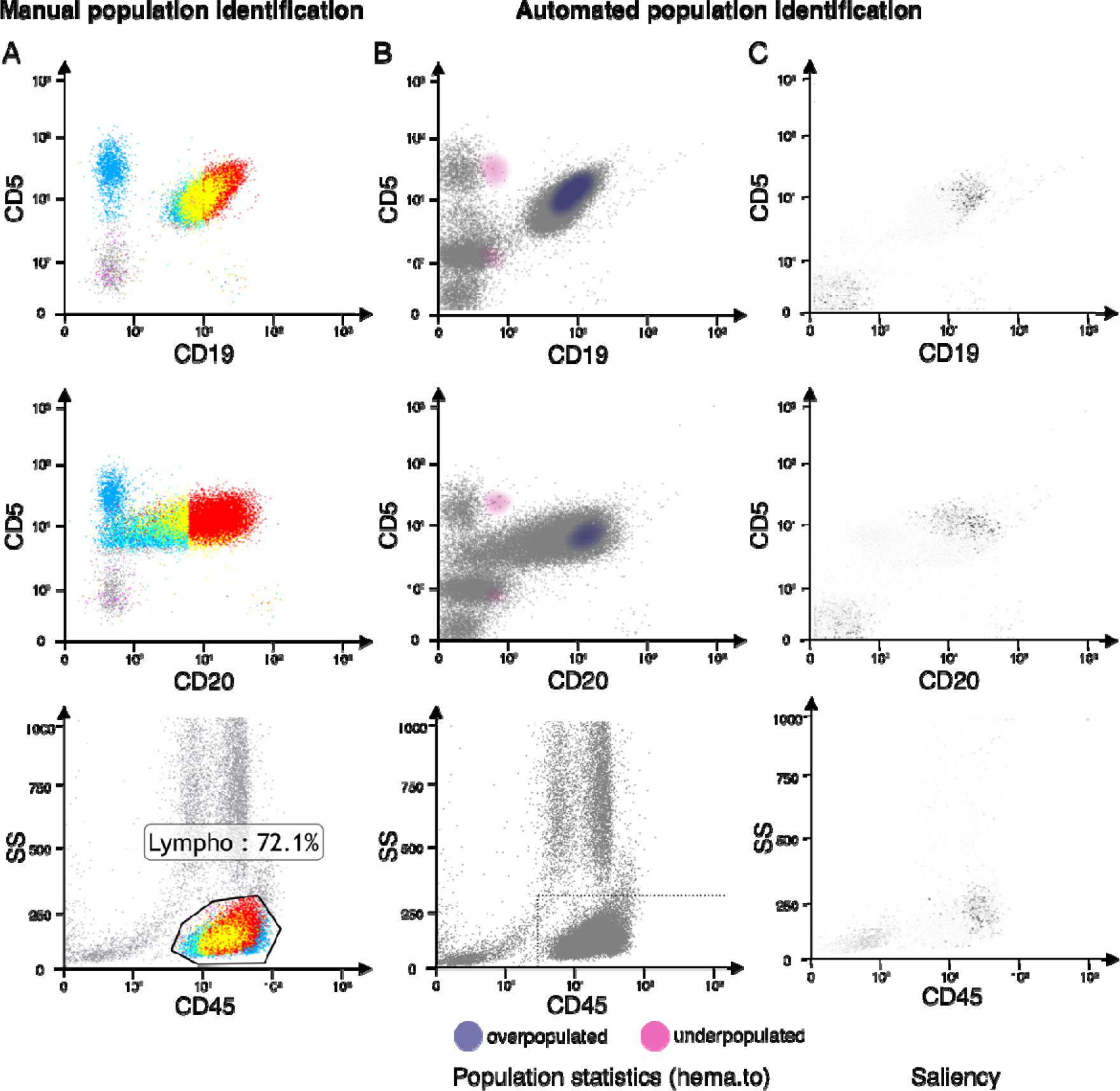
Interpretation of class predictions. Human experts identify pathogenic cell populations by a complex gating strategy over multiple 2D channel plots. The result of such a procedure is shown for a CLL sample in A). Cells are colored according to their gate properties in different scatter plots. The contrast to healthy controls is shown in B, where deviations from the average density distributions can be visualized. The test sample has a higher cell density in the same area that is also populated by manually gated cells. These are also the cells that CNN assigns the highest weight for the CLL prediction as can be seen in C). In saliency analysis the trained CNN attributes the importance a SOM node for a class label through calculation of gradients for all input channels. Based on the mapping of individual FCS events to the SOM the gradients are used to color individual events in the scatter plot (C). Darker colors represent higher gradients and thus higher importance in saliency analysis.

**Figure 4.**
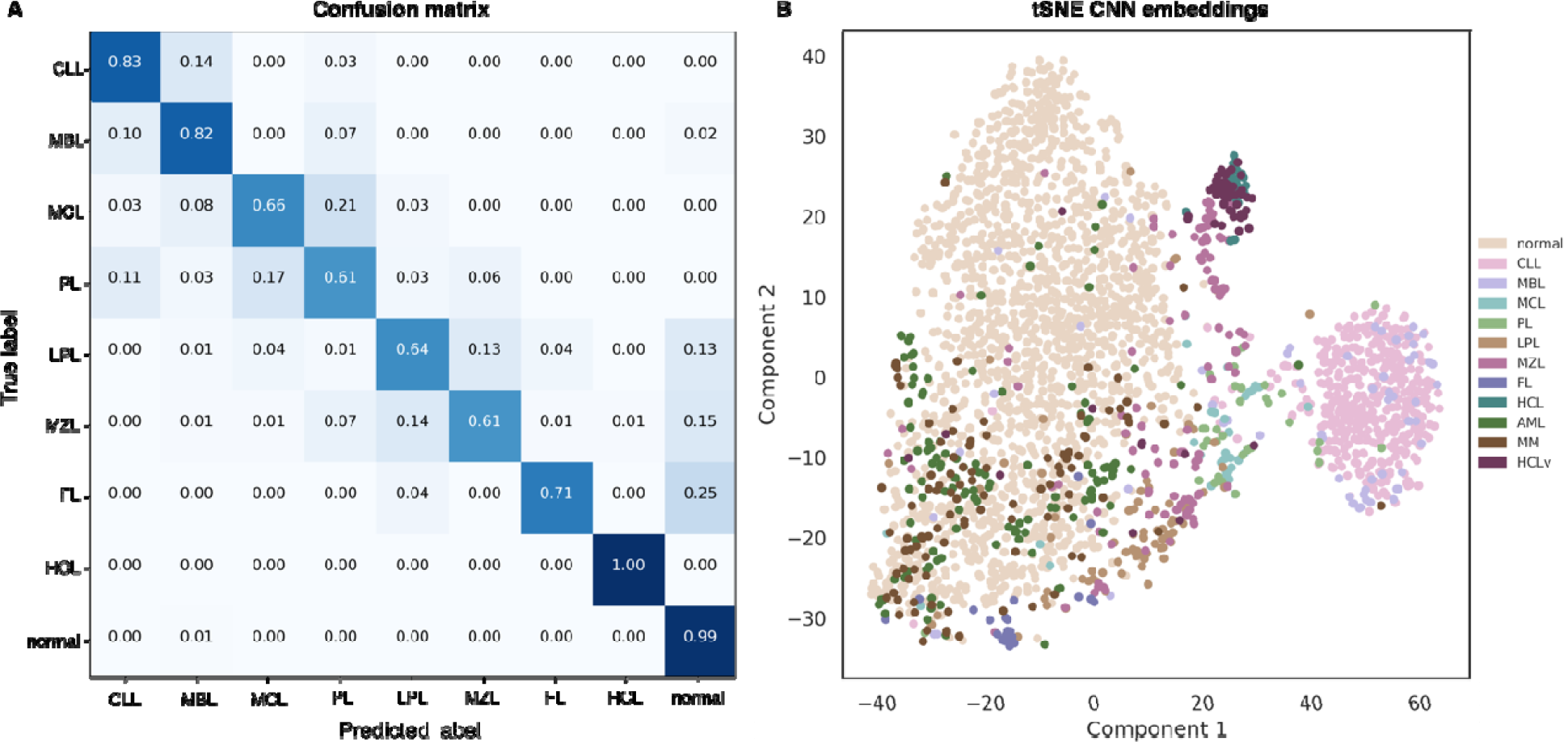
Performance of the classification process. The CNN was trained on an 9-class classification task for B-NHL subtypes (CLL, MBL, MCL, PL, LPL, MZL, FL, HCL), and normal healthy samples. The prediction accuracy was computed for a time-based holdout of 2378 cases collected after 2018-04-01, achieving a weighed F1 score of 0.92 and average F1 score of 0.73 on the 9-class problem. A) The confusion matrix also indicates the performance of the CNN for each subtype with higher error rates between clinically similar subtypes. B) This similarity can also be observed for single cases in t-SNE embeddings of the intermediary output from the concatenation layer, showing clusters of MCL/PL and MZL/LPL. Additional cases with diagnoses that the network did not learn, such as AML, MM, and HCLv are reconsidered in the discussion of the main text.

We also adapted an approach from Simonyan *et al*. and a software implementation by Kotikalapudi *et al*. (18,19) to visualize which cells were most informative for the classifier to support a certain diagnosis. All cells are assigned to nodes of the SOM and the importance or saliency of a node is defined as the maximum gradient over all input channels. SOM node saliency values were mapped back to single MFC by assigning the saliency value of the nearest SOM node to each event.

## Results

### Subtype prediction and visualization

The result of the classification process of a sample is a score for every B-NHL subtype learned by the CNN. The subtype with the highest score is the likeliest diagnosis and is used for performance readout (top-1-accuracy). In Figure 3, selected scatter plots are shown for a representative CLL sample, that has correctly been classified. The values of the other subtypes, as well as their visualizations, that is density plots and saliency maps of all scatter plots, can be reviewed at https://www.igsb.uni-bonn.de/en/research/flowcat. On the left side of Figure 3, the results of a standard manual gating strategy are shown, whereas the middle and the right side depict AI-augmented density plots and saliency maps. A likely pathological population has been identified by positivity for CD5, CD19 and CD20 after gating on lymphocytes applying CD45 and SS. A similar cell cluster yields the strongest signal for the CLL subtype as shown in the saliency analysis without prior gating. Population density shows an overpopulation in the area in comparison to healthy data.

### Performance of B-NHL classification

The top-1-accuracy rate of the classifier for the seven B-NHL subtypes plus healthy controls was computed on a hold-out test data set of 2,378 samples, yielding an average F1 score of 0.78 and a weighted F1 score of 0.94. The confusion matrix, however, indicates that misclassifications are non-uniformly distributed (Fig. 4 A). Especially the subtypes PL/MCL and MZL/LPL are more likely to be mistakenly reversed, which is plausible because of the higher similarity of their flow cytometric profiles, respectively. When distinguishing only between B-NHL and healthy control, the average and weighted F1 score of the two classes of comparable size increases to 0.98. The percentage of lymphoma cells has been estimated by human experts for each sample, with the lowest category being 0.1% lymphoma cells. As MBL is defined by the presence of less than 5000 cells with the typical CLL profile in flow cytometry, we listed MBL as a separate class in the confusion matrix. This allowed a more fine grained analysis of classification sensitivity: while there are no false negatives for CLL, some of the cases labeled as MBL were misclassified. An MBL case of the test set within the category of 0.1% lymphoma cells was classified as LPL, whereas most of the MBL cases that were misclassified as CLL had a number of lymphoma cells that was close to the distinguishing, however, arbitrary threshold of 5,000/50,000=10%. One should also consider, that the high number of MBL cases that were classified as CLL and vice versa, are a technical artifact that will decrease the F1 scores of the nine-class-problem but does not affect the F1 scores of classification problems where these subtypes are merged. The error rates in between the other B-NHL classes, however, reflect a meaningful phenotypic similarity of these disorders that can also be seen in the hierarchical clustering of the confusion matrix and the t-SNE visualization of SOM and intermediate model embeddings in Figure 4 B. Besides the aforementioned compact clusters the samples of MZL are more spread out with some closer to the CD5-positive MCL cases, which has also been reported for some cases (16).

In general, the large area under the curve of the receiver operating characteristics indicates a high classification performance (Supplementary Figure 3). Thresholds can be adjusted to arbitrary accuracies for the final model: A cutoff of 0.8 for instance did achieve a 95% accuracy on 70% of the samples of the validation and test set. This means the majority of samples are correctly classified with high confidence and the classifier could be used for technical validation.

Performance comparison between our SOM-CNN classifier and other machine learning approaches such as dense network and random forest models shows its superiority (Supplementary Figure 4). Noteworthy is also the remarkable gain in performance for a growing number of training samples that still has not reached a plateau, indicating that an even larger training set would further increase accuracy.

### Channel importance analysis

The contribution of each marker that was measured by flow cytometry to the classification accuracy can be assessed by occluding this channel and determining the difference in the performance. In principle, we expect a more considerable drop in accuracy for a subtype, when a diagnostically relevant marker is occluded. As the whole panel is already optimized for B-NHL, an occlusion test can be used to validate that our model learned the known biological differences, such as CD5 positive and negative subtypes, rather than to gain new insights. Occlusion analysis on the tube level show higher importance of tube 1 for all lymphoma groups, except HCL, which presents higher occlusion loss in tube 2 (Supplement Figure 5). Occlusion of single channels shows high importance of CD5 (tube 1) for CLL, MBL, MCL and PL, whereas CD20 (tube 1) is particularly important for differentiating MCL, PL and FL. For HCL the classification accuracy drops when CD103, CD22, CD11c, or CD25 is removed from the panel.

## Discussion

The current version of our SOM-CNN based classification method is able to differentiate eight subtypes of B-NHL and normal controls with high accuracy. The AI operates directly on compensated MFC data without the need for prior gating or manual data cleansing. For a considerable proportion of samples, that are regarded as easy to classify, the AI might thus contribute to a speed up of the diagnostic process. However, even more interesting might be the second opinion the AI can provide for difficult cases via saliency maps that point to cell clusters and suggest to which subtype this pattern would fit. This might be an additional alert that could improve the sensitivity in detecting lymphoma, before the clinician makes the final decision.

The hierarchical clustering of the confusion matrix for the multiclass problem as well as the t-SNE plots already suggest that the AI was able to learn a representation of the MFC data that mirrors our knowledge about the relatedness of the subtypes. The embedding that the network provides, can be considered as a lower-dimensional space for the MFC data, that preserves the properties we are interested in and, thus, clinically similar cases should be located close to each other. Indeed, samples with LPL/MZL and PL/MCL are part of the same clusters in the t-SNE visualizations, which is in agreement with the literature (20,21).

In general, the spatial representation of subtypes works well, if the information that was acquired with the selected markers, is sufficient for their differentiation and if these labels were used during the training process of the model. The best clustering for the subtypes we are interested in, was achieved for an embedding of the trained CNN. If limit the cluster analysis for instance to the SOMs of single tubes, the clear separation between CD5 positive and negative subtypes is lost, as can be seen in tSNE plots of tube 2 and 3 where this marker is missing (Supplemental Figure 6). In contrast, AML and MM samples that were measured with the supposedly unsuitable panel of B-NHL do form distinct clusters in the analysis of the SOMs. However, this substructure is lost when the embedding of the classifier is used, as the network was not trained on these subtypes.

This also brings us to the question how machine learning can help to optimize the marker selection in a diagnostic setting, where the number of channels is limited. Occlusion analysis of our classifier show – not surprisingly - the pivotal importance of marker CD5 by a tremendous loss of accuracy for correctly identifying CLL, MBL, MCL, and PL (Supplement Figure 5). Likewise, more subtype specific markers such as CD103 for HCL and CD10, CD20 for FL can be identified by this approach (Supplement Figure 5). It is therefore conceivable, that a set of hematologic disorders one aims to distinguish, is first screened by an extended panel of markers and the occlusion analysis are then used to select the most informative markers for this task in a high throughput setting. The accuracy can then be further increased by continuously training on as many samples as possible.

In conclusion, our work is the first application of AI for the assessment of clinical flow cytometry data, but it is just one additional piece in a long series of publications that showed how AI can increase sensitivity and specificity in health care. (22) For the further convergence of human and artificial intelligence, the key challenges for the classification of flow cytometry data will be growing the size of the training data, particularly for the rare subtypes. This will also require to adapt models in such a way, that they can handle data from in different labs. Future work should therefore also focus on the question how knowledge, that has been learned from one data set can be transferred to another. (23)

## Data Availability

All data are available upon reasonable request. Please send request to P.M.K

## Acknowledgements

M.Z. received a PhD grant from Berlin Institute of Health

## Ethics Approval

IRB or ethics approval does not apply as the study was conducted on fully anonymized retrospective patient data. Waiver was granted by the University of Bonn Medical Faculty Ethics Committee.

## Code Availability

The code is available under an open source license from the following git repository: https://github.com/xiamaz/flowCat. Additionally, we set up a web service at https://hema.to that offers a graphical user interface for processing FCS files with the CNN described in this paper. Our reported accuracy for predicting lymphoma subtypes will only be achieved if the same antibody color combinations are used (Supplemental table 1). All visualizations were implemented in matplotlib and d3.

## Data Availability

All data are available upon reasonable request. Please send request to P.M.K

